# Hybrid Video and Instance Segmentation for Surgical Scene Understanding in Robotic Mitral Valve Surgery

**DOI:** 10.64898/2026.11.16.26367936

**Authors:** Christina Waldron, Rohit Gulve, Nikhil Bhasin, Zaid Al-Shoha, Heidi Eren, Nashita Bhuiyan, Mina Zaky, Danial Ahmad, Viswajit Kandula, Sriharsha Talapeneni, Ebony Jernigan, Marc Pelletier, Nasir Rahim, Arnar Geirsson, Makoto Mori

## Abstract

**Objective:** Artificial intelligence-based surgical scene segmentation is fundamental to surgical scene understanding. However, the optimal segmentation strategy for robotic cardiac surgery remains unknown. We developed and evaluated a hybrid segmentation framework combining Mask R-CNN and Segment Anything Model 2 (SAM2) for surgical scene recognition during the entry phase of robotic mitral valve surgery.

**Methods:** Intraoperative videos from ten robotic mitral valve operations performed at two institutions were analyzed. Seven anatomical structures and surgical object classes (ascending aorta, diaphragm, lung, pericardium, right atrium, robotic arms, and sutures) were manually annotated from 2172 image frames. We created three patient-level training, validation, and test splits after ruling out any split where an object class was completely missing from validation or test. Three segmentation approaches were compared: Mask R-CNN, Mask R-CNN-prompted SAM2 for all seven classes, and a hybrid late-fusion model combining SAM2 for anatomical structures and Mask R-CNN for robotic arms and sutures. Model performance was assessed using pixel accuracy, mean intersection over union (IoU), and Dice score.

**Results:** Across the three patient-level splits, the hybrid late-fusion model achieved the highest average overall segmentation performance, with pixel accuracy of 0.79-0.91, mean IoU of 0.64-0.74, and mean Dice of 0.76-0.84. At the object level, late-fusion Dice scores across the three splits were 0.86-0.96 for aorta, 0.38-0.90 for diaphragm, 0.77-0.92 for lung, 0.48-0.77 for pericardium, 0.68-0.90 for right atrium, 0.95-0.97 for robotic arms, and 0.66-0.76 for sutures. Mask R-CNN-prompted SAM2 outperformed Mask R-CNN for the aorta in all three splits and for the lung, pericardium, diaphragm, and right atrium in two of three splits, whereas Mask R-CNN was more reliable for sutures, including Dice scores of 0.76 vs 0.21 and 0.66 vs 0.53 in Splits 2 and 3. By combining prompted SAM2 for anatomy with Mask R-CNN for robotic arms and sutures, late fusion achieved the highest mean Dice in two of three splits (0.84 and 0.76) and the highest mean Dice averaged across all three splits; all-object SAM2 propagation performed slightly better in the remaining split (0.81 vs 0.80).

**Conclusions:** A hybrid segmentation framework combining SAM2 and Mask R-CNN achieved the highest overall segmentation performance in two of three patient-level splits and performed comparably to the best-performing model in the remaining split. These findings suggest that object-specific hybrid segmentation strategies may improve surgical scene segmentation during robotic mitral valve surgery.

## Introduction

Intraoperative videos acquired during cardiac surgery remain an underutilized data source. Systematic analysis of surgical videos has the potential to characterize anatomic variation, identify opportunities to optimize operative technique, predict clinical outcomes, and serve as a powerful educational resource^1–4^. Central to these applications is the ability to accurately recognize and localize anatomic structures within the surgical field. Computer vision models, which leverage deep learning architectures, can be trained to identify anatomic structures and boundaries with high accuracy^5,6^. Although computer vision applications have been explored in cardiac surgery, little work has focused on recognition and segmentation of cardiac anatomy under direct intraoperative visualization^7^. Consequently, robust and generalizable models for this task remain underdeveloped.

Robotic cardiac surgery provides an ideal data source for developing such models. The robotic system routinely generates high-resolution videos with image stabilization, consistent camera positioning, and substantial magnification, yielding a standardized and information-rich dataset^8^. Nevertheless, the optimal approach for training object recognition models in this setting has not been established. Conventional segmentation architectures such as Mask R-CNN have demonstrated strong performance across a wide range of computer vision tasks, but they process frames independently and therefore do not explicitly leverage the temporal information contained within video sequences^9^. Temporal relationships between adjacent frames may provide additional contextual information for identifying anatomically deformable structures, such as the pericardium and right atrium, whose appearance changes with tissue manipulation and camera motion^10^. Whether a video-capable foundation segmentation model offers complementary performance to a conventional frame-wise instance segmentation model remain unknown.

Accordingly, we developed and evaluated three approaches to segmentation of anatomical structures and surgical objects during the entry phase of robotic mitral valve surgery: conventional frame-based instance segmentation using Mask R-CNN, video-based segmentation using Mask R-CNN-prompted SAM2, and a hybrid approach combining predictions from both models according to object class. We sought to compare the performance of frame-based and video-based segmentation approaches and explore whether combining their complementary capabilities could improve overall segmentation performance.

## Methods

### Study Design

This study used intraoperative video recordings from robotic mitral valve operations performed at two institutions (Yale University and Columbia University Irving Medical Center). The operations were performed on the da Vinci Xi platform. As the primary focus of the study was video analysis unrelated to clinical characteristics or outcomes, we chose 10 patients randomly from both institutions’ robotic video database. Video frames corresponding to the surgical entry phase (from chest entry to suspension of the pericardium) of the operation were analyzed. This study was approved by the Institutional Review Boards.

### Image segmentation and data preprocessing

The video segments were converted to image frames using temporal down-sampling (0.5-1 frames per second) to preserve clinically relevant transitions. Seven anatomic structures and surgical instruments were pre-defined for segmentation: ascending aorta, diaphragm, lung, pericardium, right atrium, robotic arms, and sutures. The images were annotated manually by a cardiac surgeon and trained individuals, with all annotated images reviewed for quality assurance by the cardiac surgeon (MM). The annotation process involved manually labeling the borders of each of the 7 objects of interest for every image (Supplemental Figure 1). This process yielded 2172 manually annotated images for model training, validation, and testing. Annotation data were stored in a COCO-format annotation file. Images were augmented during the training step, including multi-scale resizing (shortest edge randomly selected from 640–800 pixels), brightness adjustment (±15%), contrast adjustment (±15%), saturation adjustment (±15%), and lighting variation (PCA-based color jitter, scale factor 0.3). Horizontal flipping and rotation were not applied to avoid training the model in unrealistic orientations of the surgical scene.

### Object segmentation model development and validation

Three dataset configurations were created, each containing patient-level image sets assigned to a different training/validation/testing split (Supplemental Table 1). The split was performed at the patient level, meaning that all frames from the same patient were kept within the same subset. This was done to prevent data leakage between training and testing and to evaluate the model performance on unseen patients.

#### Mask R-CNN segmentation

Mask R-CNN was used as the baseline segmentation model. The model was implementedusing Detectron2 and used a ResNeXt-101-FPN backbone with weights initialized from a COCO-pretrained Mask R-CNN model. The final classification head was modified to predict the seven classes in the surgical dataset.

The training was performed for 15,000 iterations with a batch size of 2 and a base learning rate of 0.00025. The learning rate was reduced by a factor of 0.1 at 10,000 and 13,000 iterations, following a 1,000-iteration warmup period. Gradient clipping was enabled to improve training stability, using L2 norm clipping with a threshold of 1.0. During testing, a score threshold of 0.05 was used with a maximum of 100 detections per image, and input images were resized to a minimum of 800 pixels and a maximum of 1,333 pixels on the shortest and longest edge, respectively. After training, Mask R-CNN predictions were converted into semantic label maps. These semantic masks were later used for model performance evaluation and for the fusion model experiments.

#### Mask R-CNN and SAM2 all-object combination

An additional experiment was performed by combining Mask R-CNN and SAM2 for all objects. In this approach, Mask R-CNN predictions were used to initialize or guide SAM2 object segmentation, and SAM2 was then applied across all seven classes. The SAM2.1 Hiera Large model was used. This experiment was included to test whether applying SAM2 to every object would improve performance over Mask R-CNN alone.

However, this all-object combination was not consistently better across all metrics. Mean IoU improved over Mask R-CNN alone in all three splits, while mean Dice improved in Split 1 but decreased slightly in Splits 2 and 3. This observation led to the application of the late fusion approach below.

#### Late fusion approach

Following the observation that Mask R-CNN predicted surgical objects well while Mask R-CNN-prompted SAM2 predicted anatomical objects well, we employed a late-fusion strategy to combine the strengths of both models. This late-fusion strategy was determined post-hoc in an exploratory framework. The fusion was performed after generating semantic label maps from Mask R-CNN-prompted SAM2 and Mask R-CNN. In the final standardized approach, Mask R-CNN-prompted SAM2 was used for the 5 anatomical classes (aorta, pericardium, lung, right atrium, and diaphragm) while Mask R-CNN was used for the 2 surgical object classes (robotic arms and sutures).

For each frame, the Mask R-CNN-prompted SAM2 label map was used as the base prediction. The robot-arm and suture pixels from the SAM2 output were removed. The corresponding robot-arm and suture predictions from the Mask R-CNN semantic mask were then inserted into the final fused mask. This produced a final semantic segmentation map containing anatomical structures from Mask R-CNN-prompted SAM2 and surgical objects from Mask R-CNN.

### Model performance assessment

All methods were evaluated on the held-out test set of each dataset configuration. The evaluated methods included Mask R-CNN, Mask R-CNN-prompted SAM2 for all objects, and the final late-fusion method.

For evaluation, the predicted semantic label maps were compared with the ground-truth COCO annotations on a frame-by-frame basis. The ground-truth annotations were converted into semantic label maps before comparison. For each class, the intersection, union, predicted pixel count, and ground-truth pixel count were accumulated across all evaluated frames.

The main evaluation metrics were pixel accuracy, intersection over union (IoU), and Dice score. Pixel accuracy measured the percentage of correctly classified pixels across the full image. IoU measured the overlap between the predicted and ground-truth masks relative to their union. Dice score measured the overlap between the predicted and ground-truth masks and was used as the main class-wise metric. Class-wise IoU and Dice were calculated for all seven classes. Mean IoU and mean Dice were then computed by averaging the class-wise scores across the seven foreground classes. Dice was used as the primary metric to assess segmentation overlap for individual anatomical classes and surgical instrument classes. This allowed both overall model performance and class-wise performance to be compared across the different dataset configurations.

Classes with lower performance metrics were evaluated by visually inspecting predicted masks to qualitatively delineate potential sources of segmentation error.

## Results

### Patient characteristics

Among the 10 patients undergoing robotic mitral valve operations, the mean age was 62 years (SD 18.3 years), four patients were female (40%), and all patients underwent elective mitral valve repair (nine with indication for severe mitral regurgitation, one for myxoma of the mitral valve) (Table 1). None of the patients had prior cardiac surgery. From this cohort, we manually segmented 2172 images for analysis.

**Table 1.** Patient Characteristics.

| Characteristic | Overall, n = 10<sup>1</sup> |
|---|---|
| Age | 61.7 (18.3) |
| Sex (Female) | 4 (40%) |
| Height (cm) | 168.8 (13.3) |
| Weight (kg) | 72.5 (15.6) |
| BMI | 25.3 (3.8) |
| Diabetes mellitus | 0 (0%) |
| Asthma | 1 (10%) |
| COPD | 1 (10%) |
| Chronic kidney disease | 0 (0%) |
| Hypertension | 5 (50%) |
| Peripheral vascular disease | 0 (0%) |
| Cerebrovascular disease | 2 (20%) |
| Atrial fibrillation | 2 (20%) |
| History of cancer | 1 (10%) |
| NYHA class |  |
| I | 4 (40%) |
| II | 4 (40%) |
| III | 2 (20%) |
| IV | 0 (0%) |
| Prior cardiac surgery | 0 (0%) |
| Elective operation | 10 (100%) |
| Indication |  |
| Myxoma of mitral valve | 1 (10%) |
| Severe MR | 9 (90%) |
| CPB time (min) | 127.9 (53.6) |
| Cross-clamp time (min) | 87.0 (43.7) |

BMI: Body mass index; COPD: Chronic obstructive pulmonary disease; NYHA: New York Heart Association; MR: Mitral regurgitation; CPB: Cardiopulmonary bypass; SD: Standard deviation

### Global performance

When evaluating global model performance across all 7 classes, the late-fusion model had the highest pixel accuracy, mean IoU, and mean Dice when averaged across the three splits (Table 2). The hybrid late-fusion model had overall pixel accuracy ranging from 0.793-0.909, mean IoU ranging from 0.644-0.737, and mean Dice ranging from 0.760-0.842. For the Mask R-CNN-prompted SAM2 model, overall pixel accuracy ranged from 0.796-0.908, mean IoU ranged from 0.624-0.712, and mean Dice ranged from 0.741-0.812. For the Mask R-CNN model, overall pixel accuracy ranged from 0.786-0.871, mean IoU ranged from 0.616-0.636, and mean Dice ranged from 0.751-0.768 (Table 2).

**Table 2:**
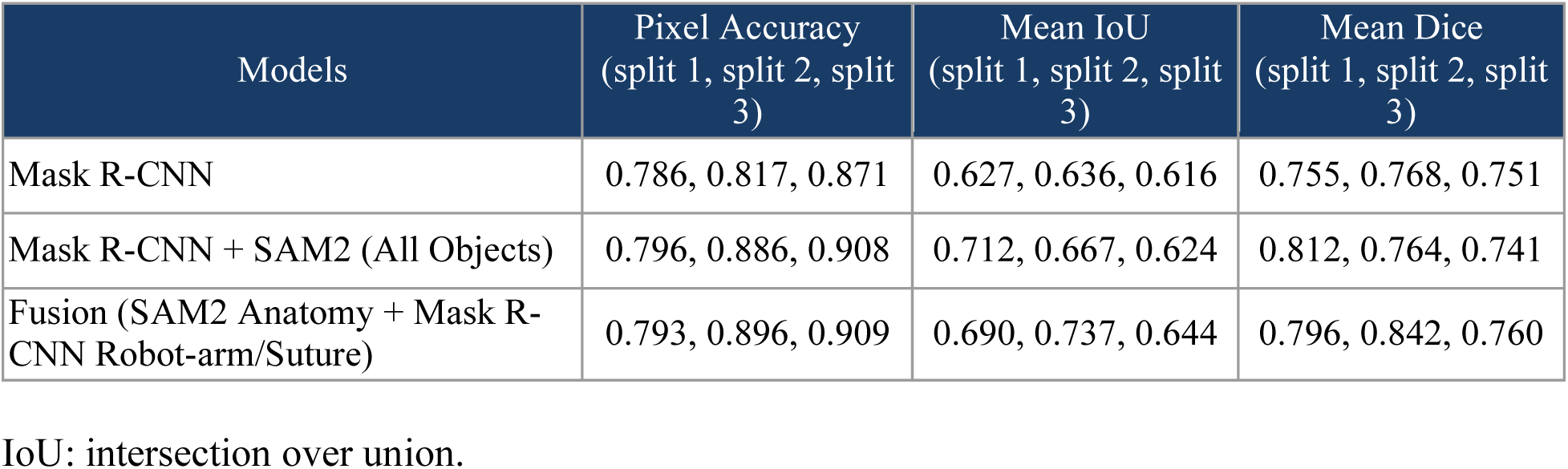
Global performance across 3 models for three data splits.

### Object-level performance

At the object level, the hybrid late-fusion model achieved the strongest overall segmentation performance when averaged across the three patient-level data splits (Table 3). The performance of the two underlying models varied across classes and splits. Mask R-CNN- prompted SAM2 performed better than Mask R-CNN on the aorta in all three splits and on the lung and pericardium in two of three splits. Dice scores for these three anatomical classes across the splits ranged from 0.481-0.959 with prompted SAM2, compared with 0.555-0.916 using Mask R-CNN. The diaphragm and right atrium also showed variable patterns: prompted SAM2 outperformed Mask R-CNN on each structure in two of three splits.

Applying SAM2 to all object classes produced variable improvements across classes and splits. The late-fusion approach, which combined prompted SAM2 predictions for anatomical structures with Mask R-CNN predictions for robotic arms and sutures, improved suture Dice compared with all-object propagation in Splits 2 and 3 but reduced it in Split 1. Consequently, the late-fusion models achieved the highest mean Dice in Splits 2 and 3 and the highest mean Dice averaged across all three splits, while all-object propagation achieved the highest mean Dice in Split 1. Neither approach was uniformly superior across the split and class.

### Visual inspection of error

Visual inspection of selected images showed that SAM2 sometimes mislabeled the pericardial edge or connecting arm as the suture, and this error was then propagated across later frames (Supplemental Figure 2).

## Discussion

In this proof-of-concept study, we evaluated three approaches to surgical scene segmentation during the entry phase of robotic mitral valve surgery: frame-based instance segmentation using Mask R-CNN, video-based segmentation using Mask R-CNN-prompted SAM2, and a class-specific hybrid approach combining predictions from both models. There were three principal findings. First, SAM2-based video propagation generally improved segmentation of anatomical structures, although performance varied across structures and patients. Second, SAM2-based video segmentation did not consistently outperform frame-based Mask R-CNN for surgical objects, particularly sutures, for which substantial performance degradation was observed in some patient-level test sets. Third, combining SAM2-based anatomical segmentation with Mask R-CNN-based surgical-object segmentation achieved the highest average segmentation performance across three patient-level splits, although it was not uniformly superior to either model alone. These findings suggest that surgical scene segmentation may benefit from combining complementary computer vision architectures rather than relying on a single segmentation approach across all object classes.

This work represents a foundation for computer vision systems capable of understanding robotic cardiac surgical scenes. Reliable recognition of both anatomy and surgical instruments is a prerequisite for higher-level applications such as operative workflow recognition, automated skill assessment, and context-aware decision support. Yet there is a lack of data demonstrating the efficacy of AI models in object segmentation for robotic cardiac operations^11^. Identifying anatomical structures from static radiographic images has been well-developed^12–15^. Computer vision models in thoracoscopic lobectomies have correlated postoperative patient outcomes with blood stains detected in the surgical field^16^. To our knowledge, this study is the first to demonstrate the feasibility of computer vision for identifying anatomical structures and surgical objects in robotic cardiac surgery.

While foundation models such as SAM2 have substantially advanced general-purpose image and video segmentation, our findings indicate that applying Mask R-CNN-prompted SAM2 propagation to all classes did not consistently provide the best performance across splits. Prior work using convolutional long-short temporal modeling suggested that temporally-aware models may have better object segmentation performance compared to static models^17^. Dynamic surgical instruments differ from native anatomy in their appearance, motion, and interaction with surrounding tissues^18^. Combining model outputs according to object type improved average segmentation performance, but did not preserve the highest-performing prediction for every class in every split. Temporal propagation may help maintain correct masks across frames, but errors in Mask R-CNN-generated prompts may also persist during propagation, potentially explaining segmentation failures in some instances. This hybrid framework provides a strategy for further investigation in surgical computer vision systems that must simultaneously interpret anatomical structures and surgical instruments.

Reliable recognition of anatomical structures and surgical objects is an essential foundation for more advanced applications of artificial intelligence in surgery. Prior surgical computer vision works have focused on laparoscopic and robotic abdominal procedures^17,19,20^. More recently, segmentation architectures such as SAM2 and Mask2Former have been investigated across natural and medical image domains, while video-based methods have enabled object propagation^21^. While such efforts in cardiac surgery has remained sparse, accurate segmentation represents only an initial component of surgical scene understanding. Recognizing the pericardium and a robotic instrument, for example, is distinct from understanding that the instrument is retracting the pericardium to expose the underlying anatomy. Progress toward context-aware surgical assistance will require integration of object recognition with temporal relationships, instrument–tissue interactions, procedural phases, and operative intent. By establishing a framework for identifying and localizing relevant structures within surgical video, our work provides an initial step toward such higher-level representations of robotic cardiac surgery.

### Limitations

This study has several limitations. First, it was conducted using videos from two institutions, but operations were performed by a single robotic mitral surgeon. Consequently, the generalizability of the proposed framework to different surgeons, institutions, and operative techniques remains to be established. Evaluation across three patient-level data splits, each containing a distinct held-out test patient, provides preliminary evidence of performance on unseen patients within this dataset, although performance varied across classes and splits.

Second, the model was developed and evaluated exclusively during the entry phase of robotic mitral valve surgery. Whether the hybrid segmentation strategy performs similarly during more technically complex portions of the operation, such as atriotomy, valve exposure, leaflet repair, and annuloplasty, requires further investigation. Third, the segmentation model was trained to recognize a limited set of anatomical structures and surgical objects selected for this phase of the operation. Expanding the model to additional anatomical landmarks, surgical instruments, and procedural steps will be necessary to enable comprehensive surgical scene understanding.

Although patient-level separation prevented frames from the same patient from appearing in both training and test sets within each configuration, each configuration contained only one held-out test patient, and training sets overlapped across configurations. In addition, the class-specific late-fusion strategy was selected post-hoc in an exploratory framework, and its performance requires confirmation on an independent dataset with the fusion rule fixed in advance. Although temporal mask propagation represents a central feature of SAM2, we did not directly quantify temporal consistency, propagation drift, or the relationship between initial prompting errors and subsequent segmentation performance. Accordingly, the proposed mechanisms underlying differences between frame-based and video-based segmentation remain inferential.

Finally, this study represents a proof of concept evaluating offline segmentation performance, and the feasibility of real-time implementation needs further evaluation.

## Conclusion

Class-specific late fusion of Mask R-CNN and Mask R-CNN-prompted SAM2 achieved the highest average segmentation performance across three patient-level splits, outperforming Mask R-CNN alone and all-object SAM2 propagation overall. These exploratory findings suggest that future surgical scene segementation may benefit from class-specific model selection, providing a foundation for further development of context-aware intraoperative AI systems.

## Data Availability

All data produced in the present study are available upon reasonable request to the authors

## Tables and Figures

**Figure 1:**
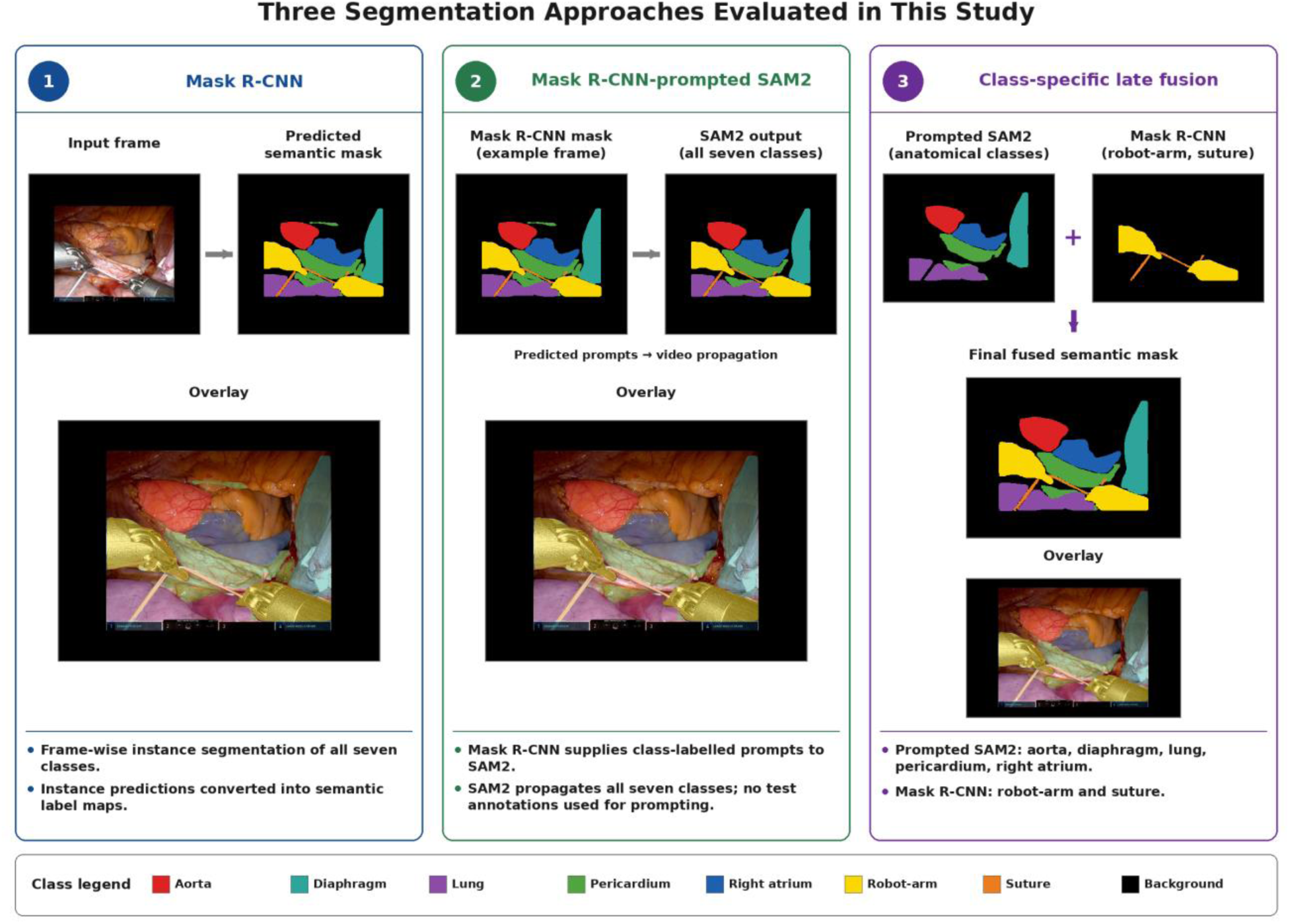
Summary of the three different modeling approaches compared. The figure summarizes the three segmentation approaches compared in this study: (1) Mask R-CNN alone, (2) Mask R-CNN-prompted SAM2 with video propagation for all seven classes, and (3) class-specific late fusion. In the late-fusion approach, robotic arms and sutures are segmented using Mask R-CNN, while the five anatomical classes—aorta, diaphragm, lung, pericardium, and right atrium—are segmented using SAM2 initialized with Mask R-CNN predictions. Test ground-truth annotations are used only for evaluation, not for prompting.

**Figure 2.**
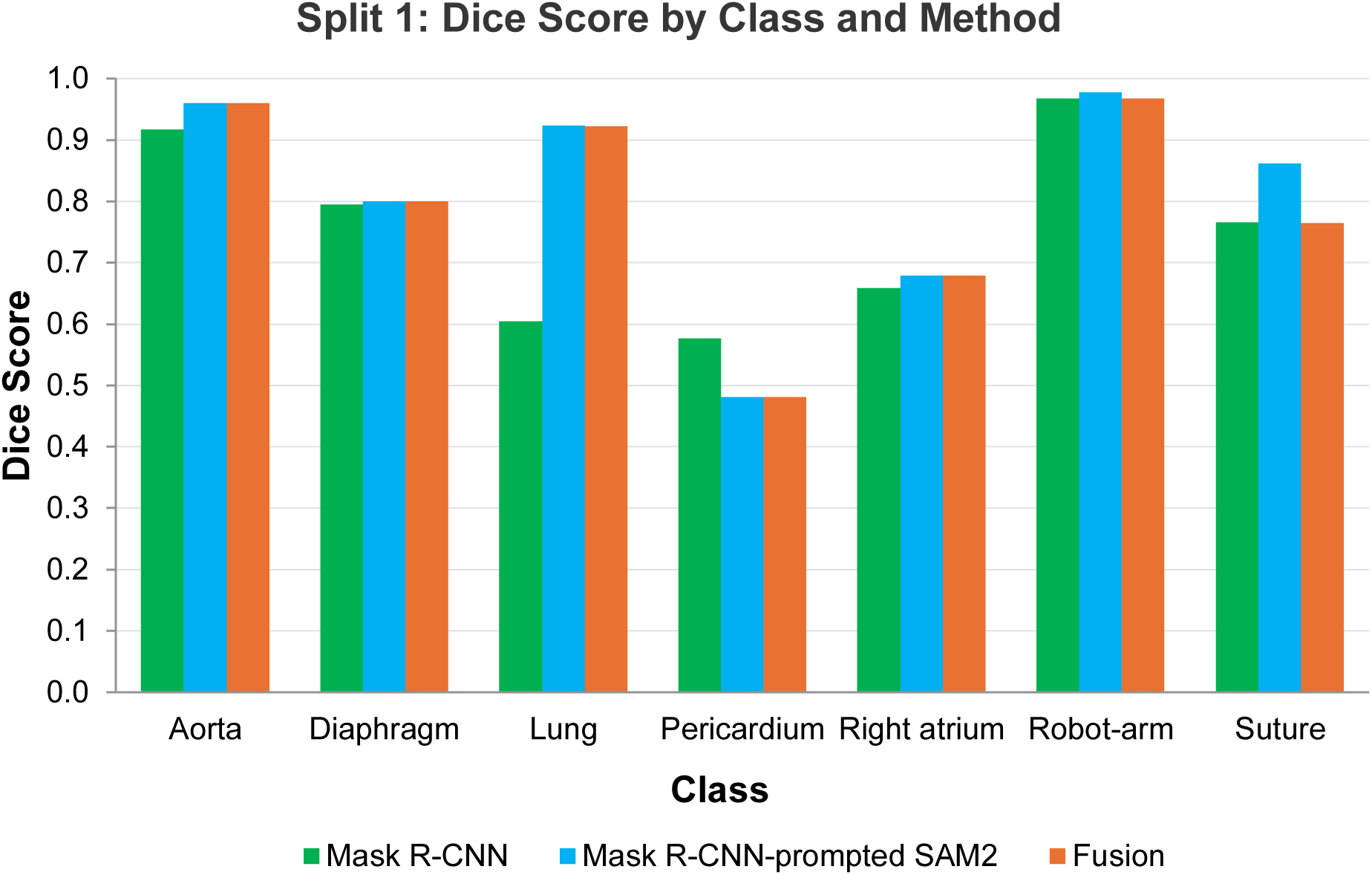
The figure summarizes object-level Dice score performance by three different models for a split (Split 1).

**Figure 3.**
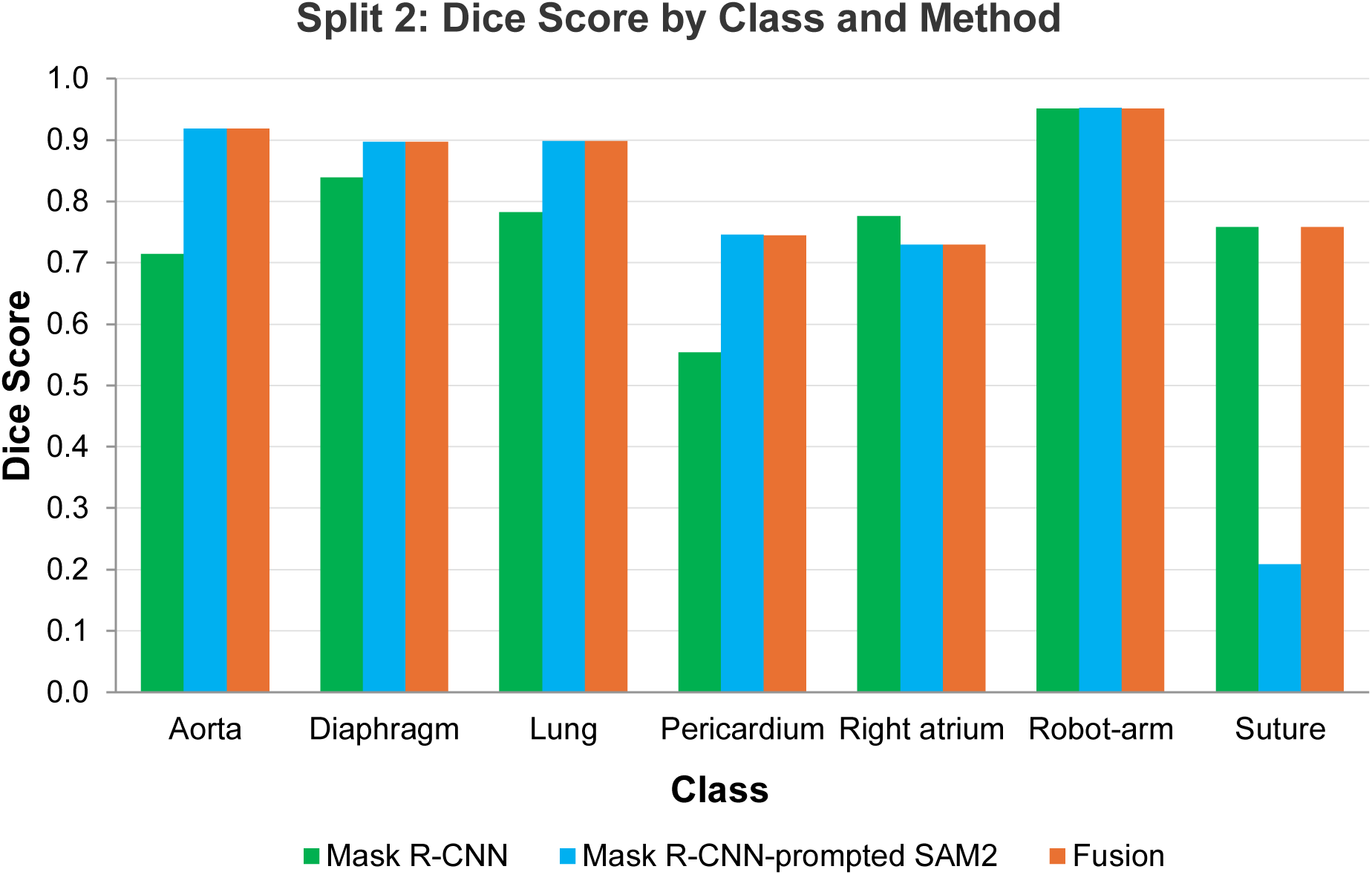
The figure summarizes object-level Dice score performance by three different models for a split (Split 2).

**Figure 4.**
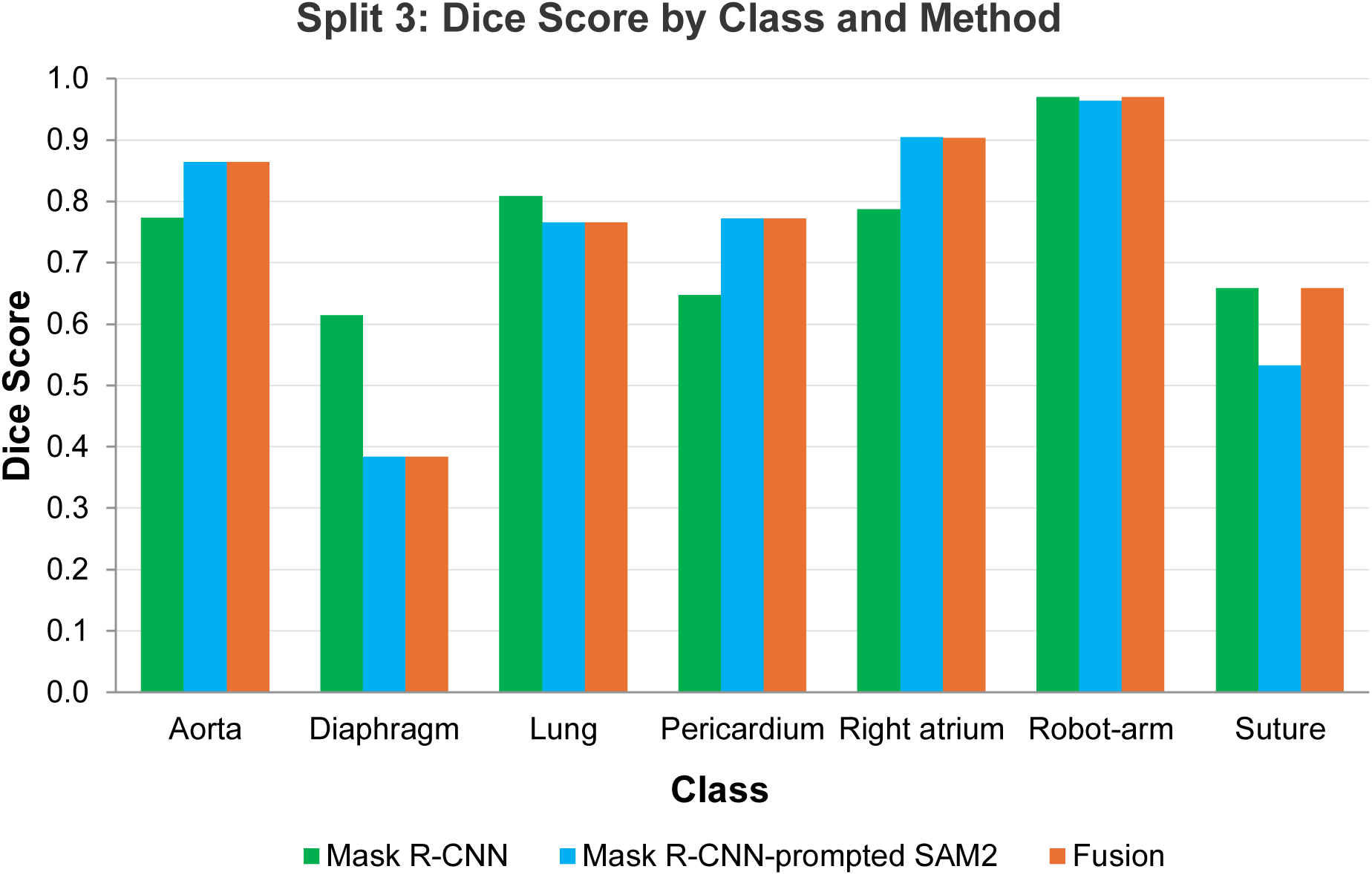
The figure summarizes object-level Dice score performance by three different models for a split (Split 3).

## Supplemental Materials

**Supplemental Table 1:** Patient data composition for each model training/validation/testing split. The table summarizes patient-level data composition within each split of model training, validation, and testing. Patient ID is a dummy variable that represents de-identified unique ID.

| Patient ID | Split 1 (test=pt8) | Split 2 (test=pt7) | Split 3 (test=pt5) |
|---|---|---|---|
| pt4 | Train | Train | Train |
| pt5 | Train | Validation | Test |
| pt6 | Train | Train | Train |
| pt7 | Validation | Test | Validation |
| pt8 | Test | Train | Train |
| pt10 | Train | Train | Train |
| pt12 | Train | Train | Train |
| pt14 | Train | Train | Train |
| pt20 | Train | Train | Train |
| pt27 | Train | Train | Train |

**Supplemental Table 2:** Split 1 Per-Class Dice.

| **Class** | **Mask R-CNN** | **Mask R-CNN-prompted SAM2** | **Leakage-free Fusion** |
|---|---|---|---|
| Aorta | 0.9165 | 0.9595 | 0.9596 |
| Diaphragm | 0.7946 | 0.7998 | 0.7998 |
| Lung | 0.6047 | 0.9231 | 0.9222 |
| Pericardium | 0.5764 | 0.4809 | 0.4809 |
| Right atrium | 0.6590 | 0.6789 | 0.6789 |
| Robot arm | 0.9677 | 0.9771 | 0.9676 |
| Suture | 0.7654 | 0.8611 | 0.7646 |

**Supplemental Table 3:** Split 2 Per-Class Dice.

| Class | Mask R-CNN | Mask R-CNN-prompted SAM2 | Leakage-free Fusion |
|---|---|---|---|
| Aorta | 0.7139 | 0.9177 | 0.9177 |
| Diaphragm | 0.8386 | 0.8972 | 0.8973 |
| Lung | 0.7820 | 0.8983 | 0.8982 |
| Pericardium | 0.5546 | 0.7451 | 0.7448 |
| Right atrium | 0.7755 | 0.7288 | 0.7288 |
| Robot arm | 0.9511 | 0.9529 | 0.9513 |
| Suture | 0.7576 | 0.2081 | 0.7576 |

**Supplemental Table 4:** Split 3 Per-Class Dice.

| **Class** | **Mask R-CNN** | **Mask R-CNN-prompted SAM2** | **Leakage-free Fusion** |
|---|---|---|---|
| Aorta | 0.7734 | 0.8641 | 0.8647 |
| Diaphragm | 0.6140 | 0.3840 | 0.3840 |
| Lung | 0.8090 | 0.7654 | 0.7654 |
| Pericardium | 0.6467 | 0.7723 | 0.7715 |
| Right atrium | 0.7877 | 0.9040 | 0.9034 |
| Robot arm | 0.9701 | 0.9631 | 0.9702 |
| Suture | 0.6591 | 0.5331 | 0.6587 |

**Supplemental Figure 1:**
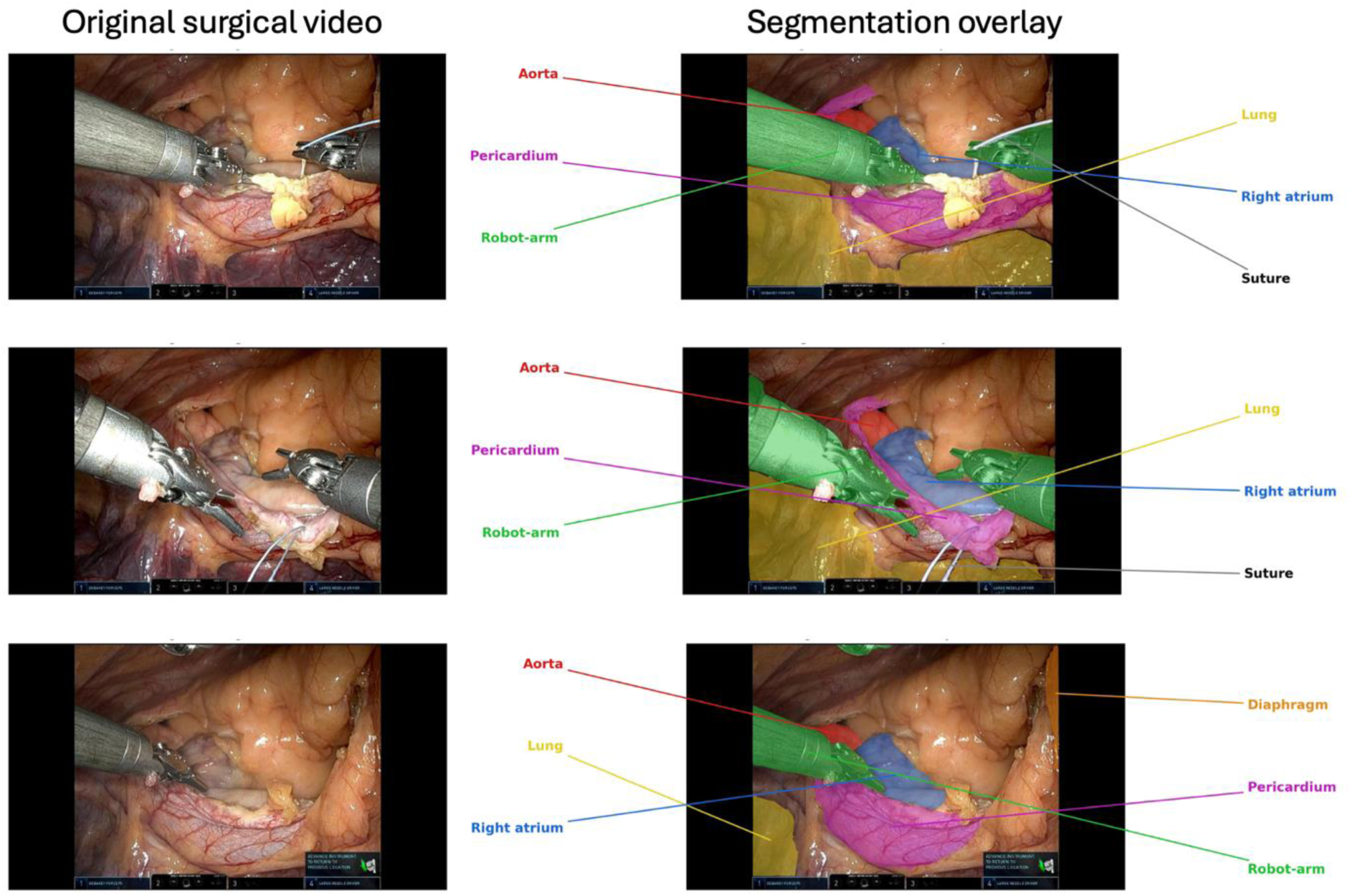
The figure shows representative surgical field views used in the analysis. The left images are the original video frames, and the right images are with segmentation overlay of objects in the field.

**Supplemental Figure 2:**
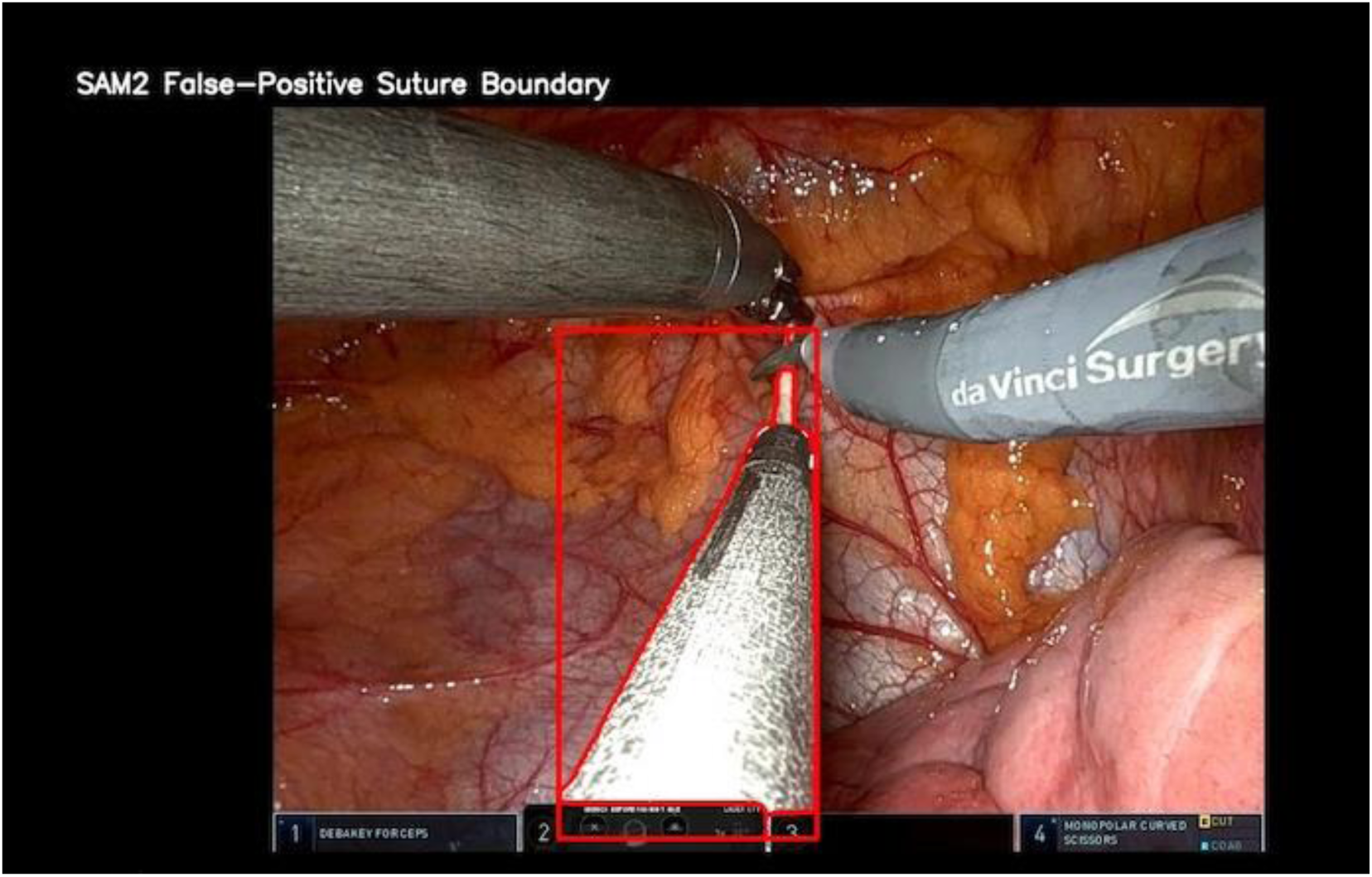
Error analysis The figure shows error in suture boundary prediction, where the model recognized the contiguous robotic arm with similar color to the suture (due to reflection) as the suture object.

